# No evidence of significant cross-reactivity between the dengue virus (DENV) and SARS-CoV-2 IgG antibodies

**DOI:** 10.1101/2021.04.19.21255725

**Authors:** Farah M. Shurrab, Fatima Humaira, Enas S. Al-Absi, Duaa W. Al-Sadeq, Hamda Qotba, Hadi. M. Yassine, Laith J. Abu-Raddad, Gheyath K. Nasrallah

## Abstract

**Background:** Several studies reported serological cross-reaction between DENV and SARS-CoV-2 IgG antibodies using rapid point of care (POC) assays. Limited data are available about cross-reactivity when testing is done using advanced chemiluminescence immunoassay (CLIA) and ELISA assays.

**Objective:** This study aims to investigate potential serological cross-reactivity between SARS-CoV-2-IgG and DENV-IgG using CLIA and ELISA assays.

**Study-design:** A total of 90 DENV-IgG-ELISA positive and 90 negative pre-pandemic sera were tested for anti-SARS-CoV-2-IgG using the automated CL-900i CLIA assay. Furthermore, a total of 91 SARS-CoV-2-IgG-CLIA positive and 91 negative post-pandemic sera were tested for anti-DENV-IgG using the Novalis ELISA assay.

**Results:** The DENV-IgG positive sera had 5 positives and 85 negatives for SARS-CoV-2-IgG. The DENV-IgG negative sera also had 5 positives and 85 negatives for SARS-CoV-2-IgG. No statistically significant difference in specificity between the DENV-IgG positive and DENV-IgG negative sera was found (p-value=1.00). The SARS-CoV-2-IgG positive sera had 43 positives, 47 negatives, and 1 equivocal for DENV-IgG. The SARS-CoV-2-IgG negative sera had 50 positives, 40 negatives, and 1 equivocal for DENV-IgG. No statistically significant difference in the proportion that is DENV-IgG positive between the SARS-CoV-2-IgG positive and SARS-CoV-2-IgG negative sera (p-value=0.58).

**Conclusions:** No evidence for cross-reactivity between the DENV and SARS-CoV-2 IgG antibodies was found.

## 1. Introduction

Severe Acute Respiratory Syndrome Coronavirus 2 (SARS-CoV-2) is a highly infectious virus causing the coronavirus disease 19 (COVID 19), which originated from Wuhan, China and has spread worldwide. However, the epidemiology and global spread of COVID-19 in high dengue virus (DENV) endemic countries shows that these regions have lower infections, transmission, and mortality rates compared to countries where DENV is not highly endemic, which were worst hit by SARS-CoV-2 infection (1). Moreover, several studies have reported serological cross-reactivity of the immune responses between SARS-CoV-2 and DENV (2, 3). These observations raised a lot of concerns regarding the possibility that pre-exposure to DENV might provide cross-protection immunity to SARS-CoV-2 infection (4). That is SARS-CoV-2 has antigenic similarity to DENV and elicits antibodies that are detected by DENV serological tests. The first reported cases of serological cross-reactivity between COVID 19 and dengue was from Singapore. These patients were dengue IgM and IgG false positive by rapid serological testing, however further testing of the original samples showed the patients were negative for DENV by RT-PCR and repeat dengue rapid test, but RT-PCR positive for SARS-COV-2 (5).

There is a limited number of studies analysing the serological cross-reactivity between SARS-CoV-2 and DENV, most of the available studies are based on the POC rapid testing kits (6-9). However, some rapid test kits have low specificity and can generate false positives (10). Hence, to investigate the potential serological cross-reactions between COVID-19 and dengue patients, we performed ELISA and CLIA automated commercial assays for detecting anti-dengue virus and anti-SARS-CoV-2 antibodies on PCR confirmed COVID-19 sera and pre-pandemic DENV positive sera, respectively.

## 2. Methods

### 2.1 Study design

A total of 91 SARS-CoV-2 IgG positive and 91 IgG negative post-pandemic sera confirmed by RT-PCR (total= 182) were available from a recent study (11). In addition, 90 DENV-IgG ELISA positive and 90 DENV-IgG negative pre-pandemic sera (total= 180) were available from a study that was conducted prior to the SARS-CoV-2 pandemic (12). The 91 SARS-CoV-2 negative sera and the 90 DENV negative sera were selected as the control group for the ELISA and the automated analyser assay testing, respectively. Specimens were collected from males, 24-69 years of age, and from nationalities of African, Asian, and Middle Eastern origins, all of whom residing currently in Qatar. This project was approved by the Institutional Review Boards at Qatar University (QU-IRB 1492-E/21).

### 2.2 Detection of DENV-IgG by ELISA

The 182 post-pandemic sera were tested for the presence of DENV-IgG using a CE-certified commercial ELISA kits (Novalisa®, dengue virus IgG; Ref. no. DENG0120, Germany). The microplate of this kit is coated with DENV virus antigens. The manufacturer reported diagnostic specificity and sensitivity of 98.0% (95% CI: 89.35%-99.95%) and 100% (95%CI: 90.75-100.0%) respectively. The detection was conducted per the manufacturer’s instructions.

### 2.3 Detection of anti-SARS-CoV-2 IgG by the CL-900i automated assay

The 180 pre-pandemic sera were tested for the presence of SARS-CoV-2-IgG using the CL-900i^®^ SARS-CoV-2 IgG (Cat. No. SARS-CoV-2 IgG121, Mindray, China) kit. The kit detects IgG specific to the spike “S” and nucleocapsid “N” proteins of SARS-CoV-2. The diagnostic specificity and sensitivity of this kit was reported earlier at 95.3% (90.1–97.8) and 90.1% (83.1–94.4), respectively (11)

## 3. Results

### 3.1 CL-900i^®^ CLIA specificity/cross reactivity with DENV-IgG

The 90 DENV-IgG positive and 90 DENV-IgG negative pre-pandemic sera samples were tested by the CL-900i^®^ SARS-CoV-2-IgG kit (Table 1). The DENV-IgG positive sera had 5 positives and 85 negatives for SARS-CoV-2-IgG. The DENV-IgG negative sera also had 5 positives and 85 negatives for SARS-CoV-2-IgG. These results indicated no statistically significant difference in specificity between the DENV-IgG positive and DENV-IgG negative p-value=1.000

**Table 1.** Outcome of SARS-CoV-2-IgG testing using the Mindray CL-900i assay on pre-pandemic sera sera that are both positive and negative for DENV-IgG.

| **Table 1.** Outcome of SARS-CoV-2-IgG testing using the Mindray CL-900i assay on pre-pandemic sera that are both positive and negative for DENV-IgG. |  |  |
| --- | --- | --- |
| **Mindray SARS-CoV-2 IgG.** | **DENV antibody positive**<br>**N (% , 95% CI)** | **DENV antibody negative**<br>**N (% , 95% CI)** |
| **Positive** | 5 (5.6%, 95% CI: 1.8-12.5) | 5 (5.6%, 95% CI: 1.8-12.5) |
| **Negative** | 85 (94.4%, 95% CI:87.5-98.2) | 85 (94.4%, 95% CI:87.5-98.2) |
| **Total** | 90 | 90 |

sera (p-value=1.00).

### 3.2 Novalisa DENV-IgG ELISA cross-reactivity with SARS-CoV-2-IgG

The 91 SARS-CoV-2-IgG positive and 91 SARS-CoV-2-IgG negative post-pandemic sera were tested by the Novalisa DENV-IgG ELISA assay (Table 2). The SARS-CoV-2-IgG positive sera had 43 positives, 47 negatives, and 1 equivocal for DENV-IgG. The SARS-CoV-2-IgG negative sera had 50 positives, 40 negatives, and 1 equivocal for DENV-IgG. These results indicated no statistically significant difference in the proportion that are DENV-IgG positive between the SARS-CoV-2-IgG positive and SARS-CoV-2-IgG negative sera (p-value=0.58).

**Table 2:** Outcome of DENV-IgG testing using the Novalisa assay on post-pandemic sera sera that are both positive and negative for SARS-CoV-2-IgG.

| **Table 2:** Outcome of DENV-IgG testing using the Novalisa assay on post-pandemic sera that are both positive and negative for SARS-CoV-2-IgG. |  |  |
| --- | --- | --- |
| **IgG**<br>**Dengue**<br>**ELISA** | **SARS-CoV-2 antibody positive**<br>**N (% , 95% CI)** | **SARS-CoV-2 antibody negative**<br>**N (% , 95% CI)** |
| **Positive** | 43 (47.3% , 95% CI: 36.7-58.0) | 50 (54.9%, 95% CI: 44.2-65.4) |
| **Negative** | 47 (51.6%, 95% CI: 40.9-62.3) | 40 (44.0%, 95% CI: 33.6-54.8) |
| **Equivocal** | 1 (1.1%, 95% CI: 0.03-6.0) | 1 (1.1%, 95% CI: 0.03-6.0) |
| **Total** | 91 | 91 |

p-value:0.580

## 4. Discussion

There has been reported cases of cross-reaction between SARS-CoV-2 and DENV and available studies have analysed this relationship mostly on rapid tests and a few on ELISA (6-9). A study analysed 55 SARS-CoV-2 positive samples on lateral flow rapid test and ELISA for dengue specifics antibodies and 95 dengue positive pre-pandemic samples on ELISA for S protein of SARS-CoV-2. The rapid tests had 12 positive cases and 14 positive cases by ELISA for dengue antibodies while the SARS-CoV-2 ELISA had 22% positive (13). However, this cross-reactivity may be assay specific and not related to antigenic cross-reaction between the dengue virus and SARS-CoV-2.

This study investigated the possible serological cross-reaction between SARS-CoV-2 and DENV. The DENV antibody positive and negative samples are pre-pandemic samples, hence eliminate the possibility of a true antigenic positive. The assay results show 5 false positives in both groups. This cross-reactivity may be assay specific, the specificity of CL-900i was reported to be 95.3% (11), this could explain the five false positive samples in both DENV groups. On the other hand, the SARS-CoV-2 antibody positive and negative groups had 43 and 50 DENV false positives, respectively and 1 equivocal each. This may be due to most patient serum used in this study were from South Asian community where it is hyperendemic to the dengue virus (14). The results between the test groups and the control groups are similar. Hence, the result is not significantly different and this in turn disapproves a theory of serological cross reaction between SARS-CoV-2 and DENV.

This is in line with a preliminary study which conducted enzyme-linked-immunosorbent-assay (ELISA) for DENV IgG-IgM on 32 anti-SARS-CoV-2 IgM/IgG positive samples and 2 COVID-IgG/IgM rapid tests on 44 DENV positive sera. The results showed 1 false positive for COVID-19 antibody in both kits, in 2 different samples among the 44 DENV positive samples and no false positives by ELISA. The study concluded there is low risk of serological cross-reaction between the 2 viruses (8). Furthermore, two studies have tested potential DENV cross reactivity with SARS-CoV-2 using automated analyser, Abbott, and Roche. Out of 46 positive dengue samples tested by Abbott and 74 testes by Roche, the results showed zero cross reactivity among the tested samples in both analysers (15, 16). These latter studies, as well as our study, supports the importance of using advanced diagnostic assays instead of rapid tests to avoid misdiagnosis, particularly in areas where DENV is endemic.

In conclusion, our results suggest that there is no strong evidence of serological cross-reactivity between SARS-CoV-2 and DENV by ELISA and CL-900i^®^ was found. Hence, the concern of cross-reactive can be downgraded among the medical community. However, it would be ideal to test for DENV and SARS-CoV-2 cross reactivity on more patient samples which are not from dengue endemic regions. In addition to that, more studies are required to analyse this cross-reaction especially using automated analysers for better strategies for the diagnosis of SARS-CoV-2 and DENV.

## Data Availability

We were constrained by our local, internal ethics policies, which entail confidentiality and therefore anonymity with regard to the samples for analysis in studies of this nature. Disclosing raw data will be breaching participant confidentiality requests to access the data should be addressed to the Ethical committee of Qatar University

## Acknowledgements

The authors would like also to thank Qatar National Library (QNL), a member of Qatar Foundation, for sponsoring the publication fees of this article. The statements made herein are solely the responsibility of the authors.

## Author contribution

Conceptualization: GKN, HMY; Methodology: FMS, FHA, DWA, ESA. Formal Analysis: FMS, FHA, ESA, LJA, GKN; Validation: GKN, FMS; Investigation: FMS, FHA, GKN, LJA, Resources: GKN, DWA, HQ; Data Curation: GKN, FMS; Writing – Original Draft Preparation: FMS, FHA, GKN, LJA; Writing, Review & Editing: GKN, HMY, LJA; Visualization: GKN, FMS; Supervision: GKN, FMS; Project Administration: GKN, DWA, Funding Acquisition: GKN, HQ. All authors have read and agreed to the published version of the manuscript.

